# Validation of the ESC 0/3h-Algorithm with a Novel High-Sensitivity Cardiac Troponin I Assay in Patients with Suspected Myocardial Infarction

**DOI:** 10.64898/2026.02.10.26346042

**Authors:** Koray Durak, Pedro Lopez-Ayala, Luca Koechlin, Jasper Boeddinghaus, Ivo Strebel, Sebastian Messingschlager, Arnaud Champetier, Emel Kaplan, Lourdes Herraiz-Recuenco, Òscar Miró, Michael Christ, Dagmar I. Keller, Javier F. Martin-Sanchez, Beata Morawiec-Migas, Jiri Parenica, Gabrielle Huré, Tom Jamie Freihofer, Karin Wildi, Paolo Bima, Luca Crisanti, Thorald Stolte, Eliska Potlukova, Danielle M. Gualandro, Felix Mahfoud, Christian Mueller, the APACE investigators

**Affiliations:** Cardiovascular Research Institute Basel (CRIB) and Department of Cardiology, University Hospital Basel, University of Basel, Switzerland; GREAT network; Department of Cardiac Surgery, University Hospital Zürich, Switzerland; BHF Centre for Cardiovascular Science, University of Edinburgh, Scotland, United Kingdom; Emergency Department, Hospital Clinic, Barcelona, Catalonia, Spain; Emergency Department, Kantonsspital Luzern, Switzerland; Emergency Department, University Hospital Zurich, Switzerland; Emergency Department, Hospital Clínico San Carlos, Madrid, Spain; Second Department of Cardiology, Faculty of Medical Sciences in Zabrze, Medical University of Silesia, Katowice, Poland; Department of Cardiology, University Hospital Brno, Brno, Czech Republic and Medical Faculty, Masaryk University, Brno, Czech Republic; Department of Intensive Care, Cantonal Hospital Aarau, Switzerland; Department of Medical Sciences, University of Turin, Turin, Italy; Department of Emergency Medicine, University of Rome, Rome, Italy; Department of Internal Medicine, Cantonal Hospital Basel-Land, Liestal, Switzerland

**Keywords:** Biomarkers, Chest Pain, Myocardial Infarction, NSTEMI

## Abstract

**Background:** There is uncertainty among physicians regarding the optimal clinical application of the novel high-sensitivity cardiac troponin I (hs-cTnI)-VITROS assay within the widely implemented upper reference limit (URL)-based ESC 0/3h-algorithm.

**Methods:** This prospective, international, multicenter study aimed to validate and compare different modifications of the URL-based ESC 0/3h-algorithm among adult patients presenting with suspected acute myocardial infarction (AMI) to the emergency department. Final diagnoses of AMI were centrally adjudicated by two independent cardiologists, blinded to hs-cTnI-VITROS concentrations. Discrimination was compared to the best validated hs-cTnT/I-assays. Clinical follow-up for cardiovascular death and AMI was conducted until five years.

**Results:** Among 1225 eligible patients (median age 63 years (IQR 52-75), 31.9% women), 18.9% were adjudicated as NSTEMI. Diagnostic discrimination of hs-cTnI-VITROS at 0h was very high (AUC 0.949, 95% CI 0.941–0.958), and comparable to hs-cTnI-Architect and hs-cTnT-Elecsys. Using the ESC 0/3h algorithm, 30.9% of patients were classified as rule-out (sensitivity 99.1%, NPV 99.5%) and 19.0% as rule-in (specificity 96.0%, PPV 82.8%). Substituting the HEART score for the GRACE score increased rule-out efficacy to 43.9%, while maintaining very high safety (sensitivity 99.6%; NPV 99.8%). Findings were consistent across predefined subgroups, in sensitivity analyses restricted to type 1 NSTEMI, using the 2h study blood sample (n=2249), and for the composite of cardiovascular death or AMI during five-year follow-up.

**Conclusions:** The ESC 0/3h-algorithm demonstrated excellent performance for rule-out of NSTEMI with greater efficacy when combined with the HEART-instead of the GRACE score.

**Registration:** https://clinicaltrials.gov/study/NCT00470587

**Clinical Perspective:** *What Is New?:*

- In a prospective, international multicenter cohort, the URL-based ESC 0/3h algorithm using the FDA-cleared hs-cTnI-VITROS assay achieved very high rule-out safety (sensitivity 99.1%, NPV 99.5%) but limited efficacy (30.9% rule-out) when combined with GRACE score <140 and symptom resolution.
- Replacing GRACE with a HEART score (<5) increased rule-out efficacy to 43.9% while maintaining excellent safety (sensitivity 99.6%, NPV 99.8%), with consistent findings across subgroups, sensitivity analyses, and long-term outcomes.

*What Are the Clinical Implications?:*

- Clinicians and health systems using hs-cTnI-VITROS can apply the ESC 0/3h algorithm with assay-conform decision values; adding clinical risk stratification is necessary to meet contemporary rule-out safety targets.

## Introduction

Acute myocardial infarction (AMI) remains the most common cause of death worldwide^1^. High-sensitivity cardiac troponin (hs-cTn) complements clinical assessment and 12-lead electrocardiograms (ECG) in the early diagnosis of patients presenting with suspected AMI to the emergency department (ED).^2–4^

Among different hs-cTn-based rapid diagnostic algorithms, the European Society of Cardiology (ESC) 0/3h-algorithm was the first to be developed and to receive a Class I guideline recommendation for the rule-out and rule-in of non–ST-segment elevation myocardial infarction (NSTEMI).^5–7^ Due to its operational simplicity, it still has great appeal to many physicians and institutions. First, in contrast to the new and possibly even faster ESC 0/1h- and 0/2h-algorithms^3,4,8–11^, the ESC 0/3h-algorithm uses the assay-specific upper reference limit (URL), that is universally highlighted in laboratory reports, as the decision value. Using laboratory reported decision values is expected to result in fewer errors as compared to the use of less familiar use-optimized cut-offs in the ESC 0/1h and ESC 0/2h-algorithm, particularly given the high fluctuation and, therefore, limited experience of residents working in the ED. Second, obtaining the second hs-cTn measurement consistently at 3h after presentation may be easier to implement in a busy ED with limited nursing staff as compared to at 1h or at 2h.^5^

The hs-cTnI-VITROS assay received FDA clearance in November 2025, and has demonstrated excellent diagnostic safety and efficacy when applied within ESC 0/1h- and 0/2h-algorithms.^12–14^ However, its performance within the widely implemented URL-based ESC 0/3h-algorithm has not been evaluated, representing a critical and unmet clinical need. Therefore, this secondary analysis of a large prospective international study aimed to I) evaluate the diagnostic and prognostic performance of the ESC 0/3h-algorithm using hs-cTnI-VITROS, II) assess the diagnostic performance of a simplified version of the ESC 0/3h-algorithm including only hs-cTnI-VITROS, but not the two additional criteria of the ESC 0/3h-algorithm (pain-free and GRACE-score<140), and III) evaluate the diagnostic accuracy of the ESC 0/3h-algorithm using an 0/3h absolute change criteria and the HEART score for ruling out patients as an alternative to the two other criteria.

## Methods

### Study design and population

This study was a secondary analysis of the Advantageous Predictors of Acute Coronary Syndromes Evaluation (APACE, ClinicalTrials.gov Identifier: NCT00470587) study, a prospective, multicenter, international diagnostic study conducted in 12 centers across five European countries^13,15–17^. The APACE study had the prespecified aim to investigate the diagnostic performance of new biomarkers, including hs-cTn assays, in adult patients presenting with symptoms suggestive of AMI to the ED.

For this analysis, exclusion criteria were 1) the presence of ST-segment elevation MI, 2) end-stage renal disease requiring long-term dialysis, or 3) an unclear final diagnosis even after final adjudication and at least one elevated hs-cTnI measurement, thereby possibly indicating AMI. Patients were further excluded if they did not have a 0h and a 3h hs-cTnI-VITROS measurement. The study was performed in accordance with the Declaration of Helsinki and was approved by the local ethics committees of all participating institutions. Written informed consent was obtained from all patients. Reporting adhered to the Standards for Reporting of Diagnostic Accuracy Studies (STARD) statement^18^ (**Supplementary Table 1**). Because of the sensitive nature of the data collected for this study, data and materials are not publicly available.

### Central adjudication of final diagnosis

Central adjudication of the final diagnosis was performed by two independent cardiologists applying current guidelines and the Fourth Universal Definition of MI.^3,4,19^ In cases of disagreement, a third senior cardiologist was consulted to reach a final decision. The adjudication process followed established methodologies, integrating both clinical and study-specific assessments, as previously described.^12,15,20^

### Hs-cTnI assay

Blood samples were collected into heparin plasma and serum tubes at presentation to the ED (0h) and serially thereafter (1h, 2h, 3h, and 6h after presentation). Serial sampling was discontinued if the patient was discharged or transferred to the catheterization laboratory for acute intervention. After centrifugation, samples were frozen at −80°C until assayed in a blinded fashion in a dedicated core laboratory. According to the manufacturer, the URL of hs-cTnI-VITROS in serum was 11 ng/L overall, with sex-specific values of 9 ng/L for females and 12 ng/L for males, and a corresponding coefficient of variation (CV) of <7% at the URL, a limit of detection of 0.39 ng/L, a limit of quantification (LOQ, <20%CV) of 1.23 ng/L (translating into a reporting limit of 1.5ng/L in clinical practice in most countries), and within-lab precision <10% at 1.84 ng/L.^21^

### Study Endpoints

The primary diagnostic endpoint was NSTEMI (type 1 or type 2) at presentation to the ED. The primary prognostic endpoint was 5-year all-cause death. The secondary prognostic endpoint was 5-years cardiovascular death or MI. The best validated hs-cTnI-assay (Architect) and hs-cTnT-assay (Elecsys Gen 5) were used for the direct comparison of the diagnostic discrimination of the hs-cTnI-VITROS assays.^13,15–17^ To assess the robustness of the diagnostic performance of the algorithms, a sensitivity analysis using only type 1 NSTEMI as outcome was performed.

### Follow-up

To assess death or major adverse cardiovascular events (MACE), follow-up was conducted at 3, 12, 24, and 60 months after discharge through telephone contact or written correspondence. The cause of death was determined using hospital records, family physician documentation, or national death registries.

### Clinical risk scores

The GRACE score was calculated using standard criteria.^3–5,22,23^ All variables were prospectively collected except for Killip class, which was assigned retrospectively based on clinical examination findings. The HEART score was calculated according to its original definition using prospectively collected data on history, electrocardiographic findings, age, cardiovascular risk factors, and hs-cTn concentrations.^24,25^

### ESC 0/3h-algorithms

For the evaluation of rule-in and rule-out strategies, the URL-based ESC 0/3h-algorithm from the 2011 and 2015 ESC Guidelines on the management of acute coronary syndromes were applied.^5^ The ESC 0/3h-algorithm uses hs-cTn measurements at ED presentation and after 3h (±30 minutes). Three different triage algorithms were evaluated:

First, the complete ESC 0/3h-algorithm using hs-cTnI-VITROS in combination with the pain-free criteria and a GRACE score <140 for rule-out. Early rule-out was defined as a hs-cTnI concentration below the URL in patients presenting >6h after chest pain onset; in patients presenting ≤6h after symptom onset, two consecutive hs-cTnI measurements below the URL were required. In all cases, rule-out additionally required resolution of symptoms prior to ED presentation and a GRACE score <140. Early rule-in was defined by markedly elevated initial hs-cTnI concentrations (≥5×URL) or by an absolute increase ≥URL on the second measurement. Additionally, the same algorithm was evaluated using an alternative and meanwhile even better validated clinical risk stratification tool, the HEART score, requiring a cut-off <5 points for rule-out irrespective of resolution of symptoms.^24–26^ As the HEART score with a cut-off <5 points has not been formally validated within the ESC 0/3h algorithm before, we additionally assessed its performance in combination with two further hs-cTn assays, hs-cTnI-Architect and hs-cTnT-Elecsys Gen-5, using previously published URL values (26 ng/L and 14 ng/L, respectively).^27–29^

Second, a simplified version of the ESC algorithm using only the hs-cTnI-VITROS criteria. Early rule-out was based on a hs-cTnI measurement below the URL, if chest pain onset had occurred more than 6h prior to ED arrival. Early rule-in was defined by markedly elevated initial hs-cTnI concentrations (≥5×URL) or by an absolute increase ≥URL on the second measurement.

Third, the ESC 0/3h-algorithm using LOQ-based criteria for the absolute change between two serial measurements of hs-cTnI-VITROS. Two consecutive hs-cTnI measurements below the URL were required in combination with an absolute change <1.5ng/L. Early rule-in was defined by markedly elevated initial hs-cTnI concentrations (≥5×URL) or by an absolute increase ≥URL at second measurement. In sensitivity analyses, alternative delta criteria were evaluated for rule-out, including a LOQ-based relative change threshold of <20% and a required absolute change of 0 ng/L.

In accordance with previous studies and ESC guidelines, target sensitivity and negative predictive value (NPV) for rule-out were >99.0% and 99.5%, respectively. Target positive predictive value (PPV) was >70%.^3,12–14^

### Statistical analysis

Continuous variables are reported as median (25^th^-75^th^ percentiles [Q1-Q3]) and categorical variables as count (percentage). Non-parametric statistical descriptors were used by default to minimize assumptions about the underlying distribution of the data. The 95% confidence intervals (CIs) were calculated using Wilson’s method if not otherwise specified. For the direct comparison of the diagnostic accuracy between the hs-cTnI-VITROS and the hs-cTnI-Architect assays, receiver-operating characteristics (ROC) curves were constructed and their corresponding area under the curve (AUC) assessed. Confidence intervals of AUC and P-values for comparison of AUCs were computed as recommended by DeLong.^30^

Safety of the ESC 0/3h-algorithms was assessed as the sensitivity and NPV of ruling out index NSTEMI and accuracy as the specificity and PPV of ruling in index NSTEMI. Efficacy was determined by the number of patients triaged towards rule-out or rule-in as a proportion of the total study sample. Additionally, diagnostic performance was assessed in I) a sensitivity analysis with NSTEMI type I as diagnostic endpoint and II) 5 predefined subgroups: age (<65 or ≥ 65 years), sex, early presenter (defined as chest pain onset ≤6h), history of diabetes, and ischemic signs on ECG (T-wave depression or ST-segment depression). For the survival analysis, patients were grouped according to their final allocation of the ESC algorithm (rule-in, observe, rule-out). Five-year all-cause mortality was plotted in Kaplan Meier curves, and the log-rank test was used for comparison between groups. For the 5-year MI or cardiovascular death endpoint, the Cumulative Incidence Function method was used for plotting the incidence of the occurrence of the composite outcome while taking competing risk (non-cardiovascular death) into account, and the Gray’s test was used for comparison between groups. Patients were included only once (first admission). In a sensitivity analysis, the different ESC 0/3h-algorithms were applied using the 2h (±30 minutes) study sample rather than the 3h study sample, as the proportion of APACE patients with missing 2h sample is lower as compared to missing 3h sample, reducing a possible biased introduced by differences in patient characteristics in patients with missing 2h or 3h study blood samples. All statistical comparisons were two-sided, and p-values of <0.05 indicated statistically significant differences. All statistical analyses were performed using R 4.4.3 (R Foundation for Statistical Computing, Vienna, Austria) and packages (with version number) are listed in the **Supplementary Table 2**.

## Results

### Baseline characteristics

A total of 1225 patients were included for validation of the ESC-0/3h algorithm in the main analysis and 2249 patients for the sensitivity analysis using the 2h-study sample (**Supplementary Figure 1A and 1B**). Patients in the main analysis had a median age of 63 (IQR: 52-75) years, 31.9% (391) were female and 54.8% (671) were early presenters (**Table 1)**.

**Table 1.**
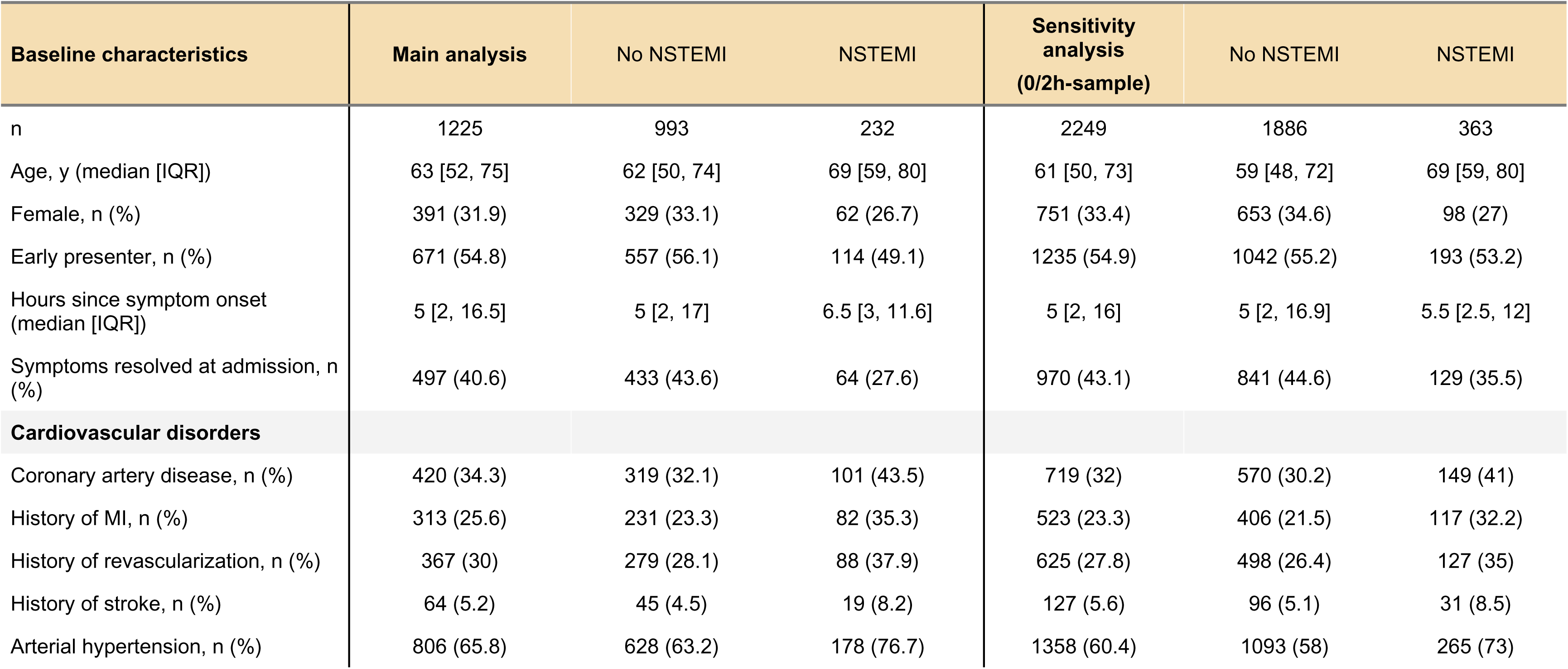
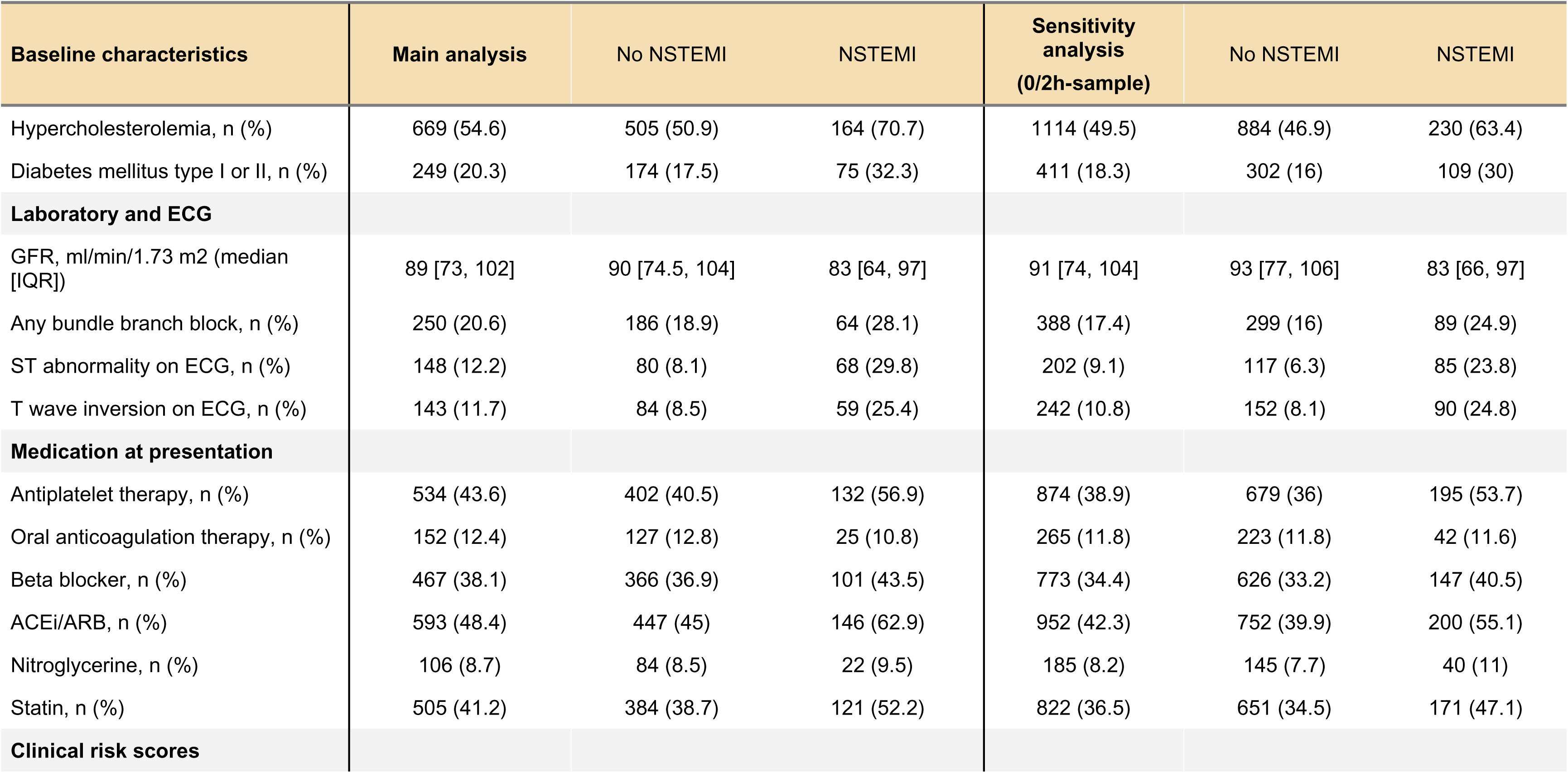
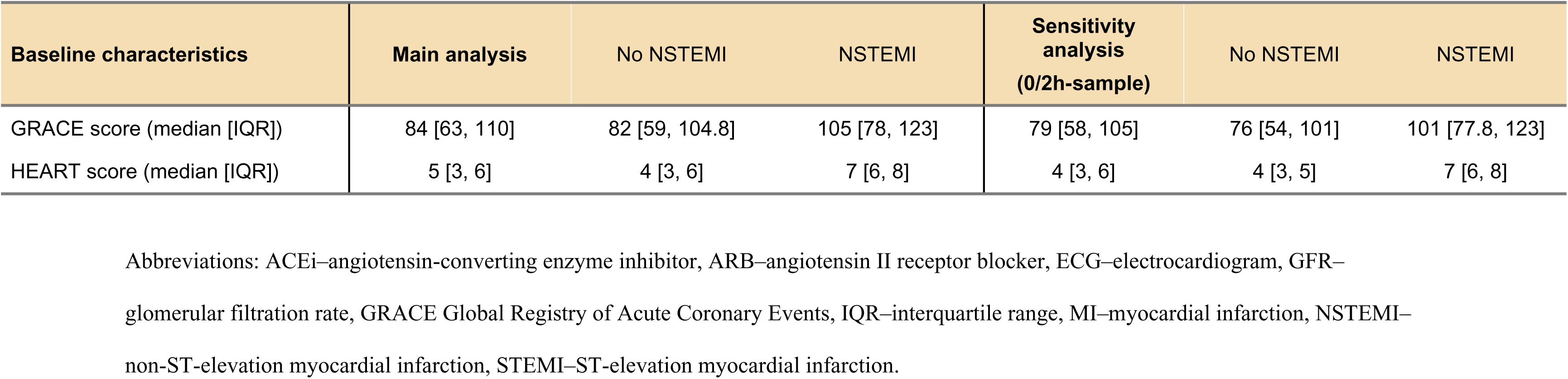
Baseline characteristics.

Concentrations of hs-cTnI-VITROS at ED presentation were higher in patients with NSTEMI than in those with other final diagnoses (P < 0.001). Median concentrations of hs-cTnI-VITROS were 68.2 ng/L (IQR: 16.4–500) in NSTEMI, compared with 3.28 ng/L (IQR: 1.58–6.45) in unstable angina, 4.07 ng/L (IQR, 1.68–12.2) in cardiac disease without coronary artery disease, 1.28 ng/L (IQR: 0.71–2.65) in non-cardiac disease, and 2.22 ng/L (IQR: 1.38–3.77; **Supplementary Figure 2**) in patients with symptoms of unknown origin.

### Diagnostic discrimination for NSTEMI

The diagnostic discrimination of hs-cTnI-VITROS at 0h was 0.949 (95%CI: 0.941–0.958) and comparable to hs-cTnI-Architect and hs-cTnT-Elecsys-Gen-5 at 0.942 (95%CI: 0.934–0.950) and 0.933 (95%CI: 0.923–0.942), respectively (**Figure 1**).

**Figure 1.**
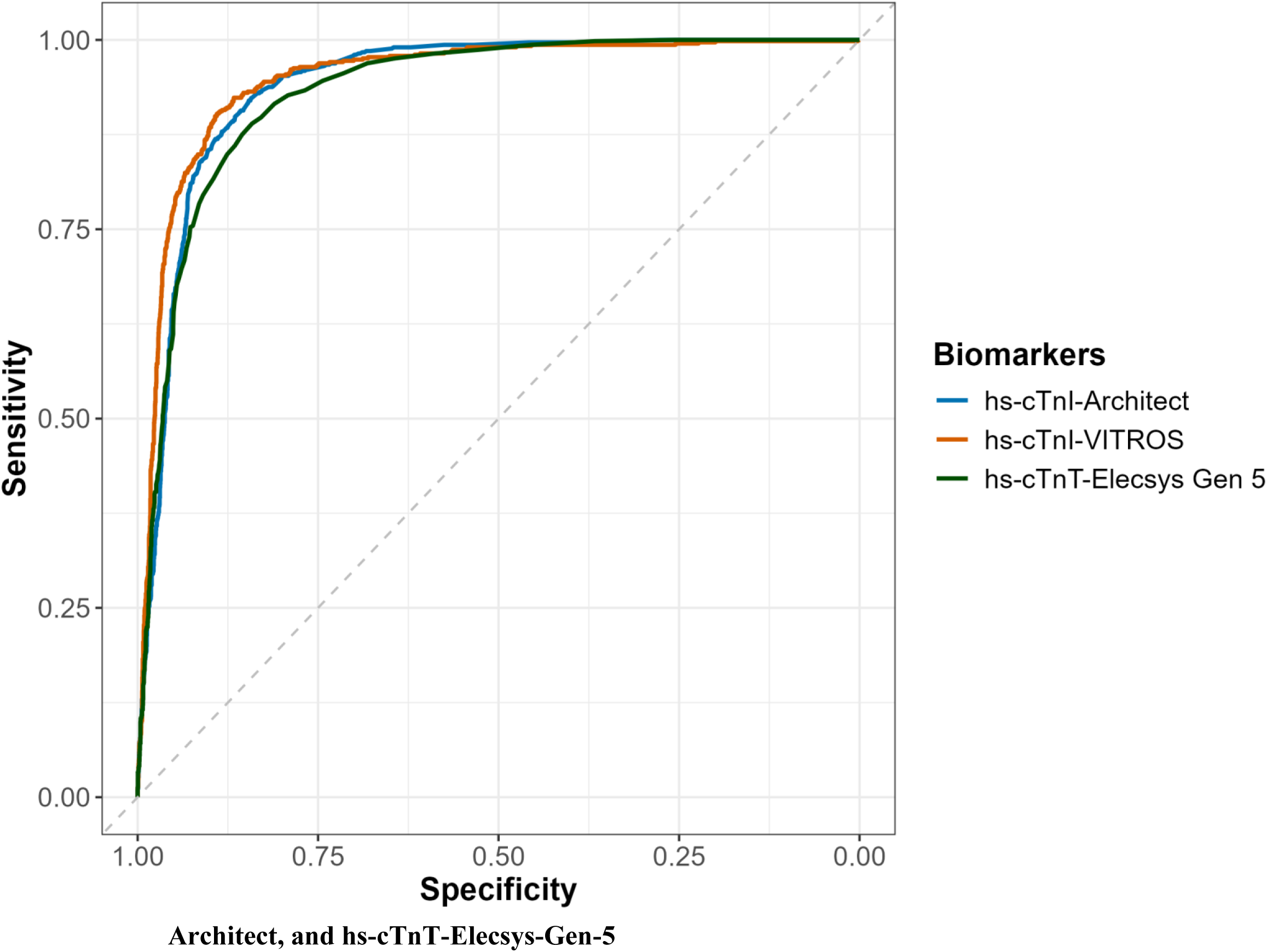
Diagnostic discrimination for NSTEMI of hs-cTnI-VITROS, hs-cTnI-Architect, and hs-cTnT-Elecsys-Gen-5: The AUC showing the discrimination of hs-cTnI-VITROS and hs-cTnI-Architect. Abbreviations: AUC– area under the receiver operating characteristic curve, hs-cTnI–high-sensitivity cardiac troponin I.

### Validation of complete ESC 0/3h-algorithms using hs-cTnI-VITROS and GRACE score

In the main cohort (n=1225; 18.9% with NSTEMI), a URL-based cutoff of <11 ng/L (applied at both 0h and 3h, or only at 0h in patients presenting > 6h after chest pain onset [direct rule-out], in combination with a GRACE score <140 and resolution of symptoms) classified 378 patients (30.9%) as rule-out, resulting in a sensitivity of 99.1% (95%CI: 96.9–99.8) and a NPV of 99.5% (95%CI: 98.1–99.9). The URL-based rule-in criteria for NSTEMI (hs-cTnI ≥55 ng/L or an absolute 0/3h change ≥11 ng/L) ruled-in 233 patients (19%) with a specificity of 96% (95%CI: 94.6–97.0) and a PPV of 82.8% (95%CI: 77.5–87.1). A total of 614 patients (50.1%) remained in the observe zone, of whom 37 patients (6%) had an adjudicated NSTEMI (**Figure 2A**). The diagnostic performance was consistent in the sensitivity analysis with NSTEMI type I and all predefined subgroups (**Supplementary Figure 3A and 4**). Similar results were obtained in the sensitivity analysis using the 2h-study sample (**Figure 2B; Supplementary Figure 3B and 4**).

**Figure 2.**
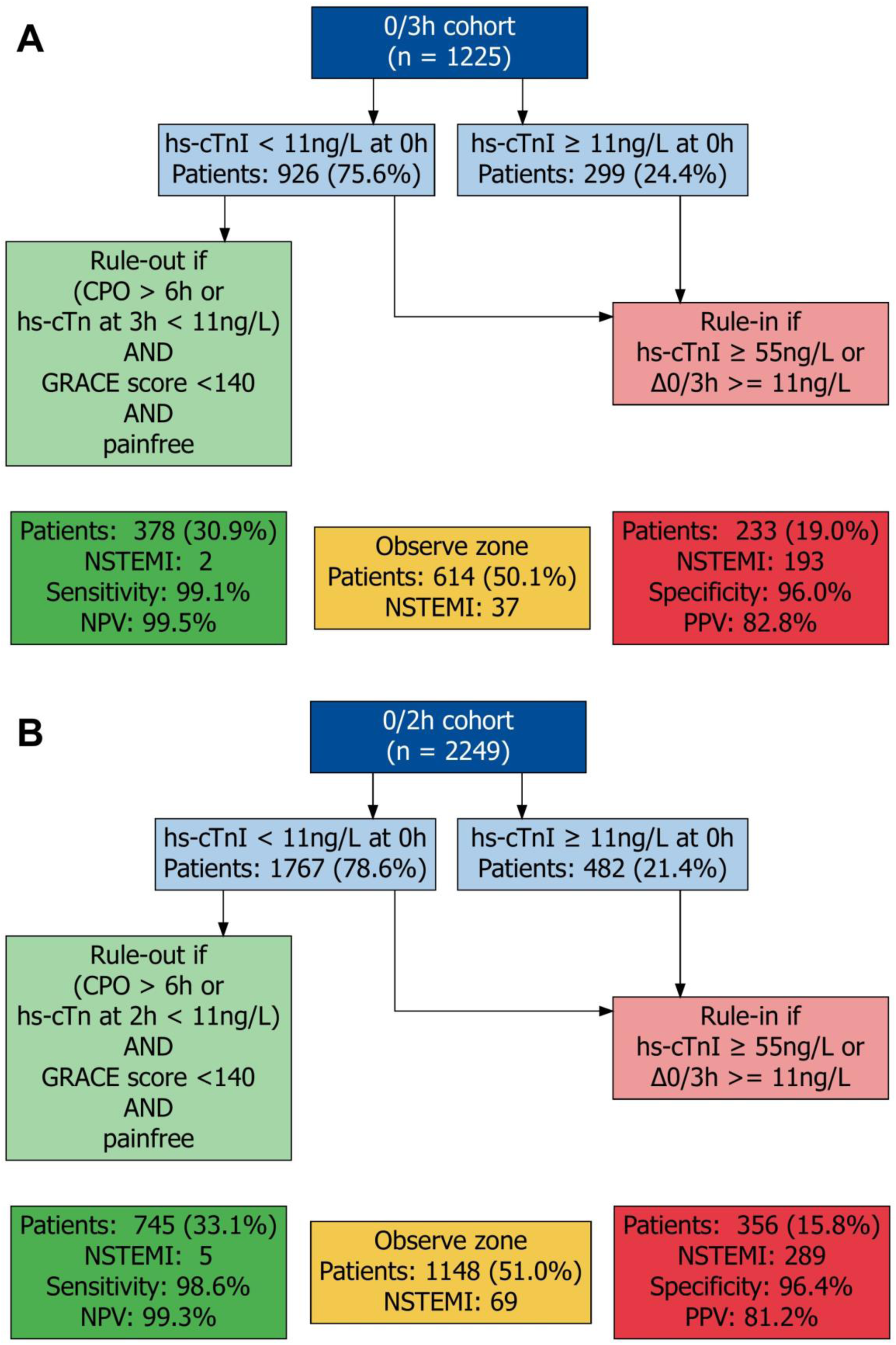
Validation of the complete ESC 0/3h-algorithm using hs-cTnI-VITROS and GRACE score. Validation of the ESC 0/3-hour algorithm using hs-cTnI-VITROS with GRACE score–based clinical risk stratification and symptom resolution in the (A) 0/3-hour cohort and (B) 0/2-hour cohort. The algorithm displays patient flow and diagnostic safety and efficacy for rule-out, observe, and rule-in categories. Abbreviations: hs-cTnI–high-sensitivity cardiac troponin I, GRACE–Global Registry of Acute Coronary Events, NPV–negative predictive value, NSTEMI–non-ST-elevation myocardial infarction, PPV–positive predictive value, Sens–sensitivity, Spec–specificity, URL–upper reference limit.

### Prognosis of complete ESC 0/3h-algorithms using hs-cTnI-VITROS and GRACE score

In the main cohort, death from any cause through 5 years occurred in 162 patients, with cumulative mortality rates of 7.5% (27 deaths) in the rule-out group, 11.9% (68 deaths) in the observe zone, and 29.6% (67 deaths) in the rule-in group (**Figure 3A**). The 5-year cumulative incidence for MI or CV death was 5.8% (95%CI: 3.7–8.7%) in the rule-out group, 9.8% (95%CI: 7.5–13%) in the observe zone, and 27% (95%CI: 21–33%) in the rule-in group (**Supplementary Figure 6A**). Sensitivity analysis using the 2h study blood sample revealed similar findings (**Figure 3B and Supplementary Figure 6B**).

**Figure 3.**
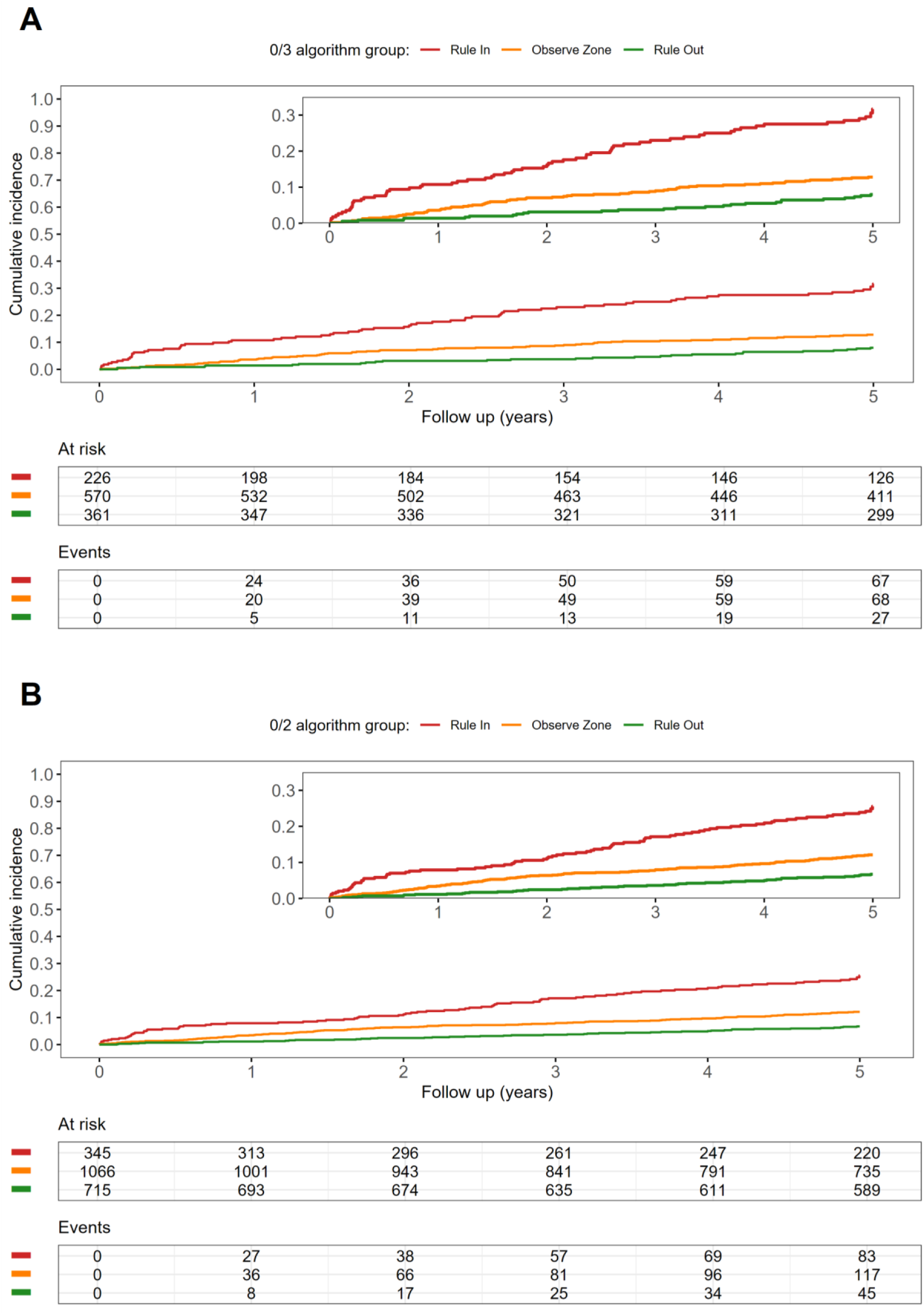
Prognosis of the complete ESC 0/3h-algorithm using hs-cTnI-VITROS and GRACE score. Kaplan–Meier plots depicting 5-year cumulative incidence of all-cause death in patients stratified by the complete ESC 0/3-hour algorithm using hs-cTnI-VITROS and GRACE score in the (A) 0/3-hour cohort and (B) 0/2-hour cohort. Log-rank test for between-group comparison for all-cause death, p<0.001.

### Validation of complete ESC 0/3h-algorithms using hs-cTnI-VITROS and HEART score

In the main 0/3h cohort (n=1225; 18.9% with NSTEMI), a URL-based cutoff of <11 ng/L (applied at both 0h and 3h, or only at 0h in patients presenting > 6h after chest pain onset [direct rule-out], in combination with a HEART score <5) classified 538 patients (43.9%) as rule-out, resulting in a sensitivity of 99.6% (95%CI: 97.6–99.9) and a NPV of 99.8% (95%CI: 99.0–100). The URL-based rule-in criteria for NSTEMI (hs-cTnI ≥55 ng/L or an absolute 0/3h change ≥11 ng/L) ruled-in 232 patients (18.9%) with a specificity of 96.1% (95%CI: 94.7–97.1) and a PPV of 83.2% (95%CI: 77.8–87.5). A total of 455 patients (37.1%) remained in the observe zone, of whom 38 patients (8.4%) had an adjudicated NSTEMI (**Figure 4A**). The diagnostic performance was consistent in the sensitivity analysis with NSTEMI type I and all predefined subgroups (**Supplementary Figure 7A and 8**). Sensitivity analysis using the 2h study blood sample revealed similar findings (**Figure 4B; Supplementary Figure 7B and 9**). Additionally, we found consistent results for the use of hs-cTnI-Architect and hs-cTnT-Elecsys-Gen-5 instead of hs-cTnI-VITROS (**Supplementary Figure 10 and 11**).

**Figure 4.**
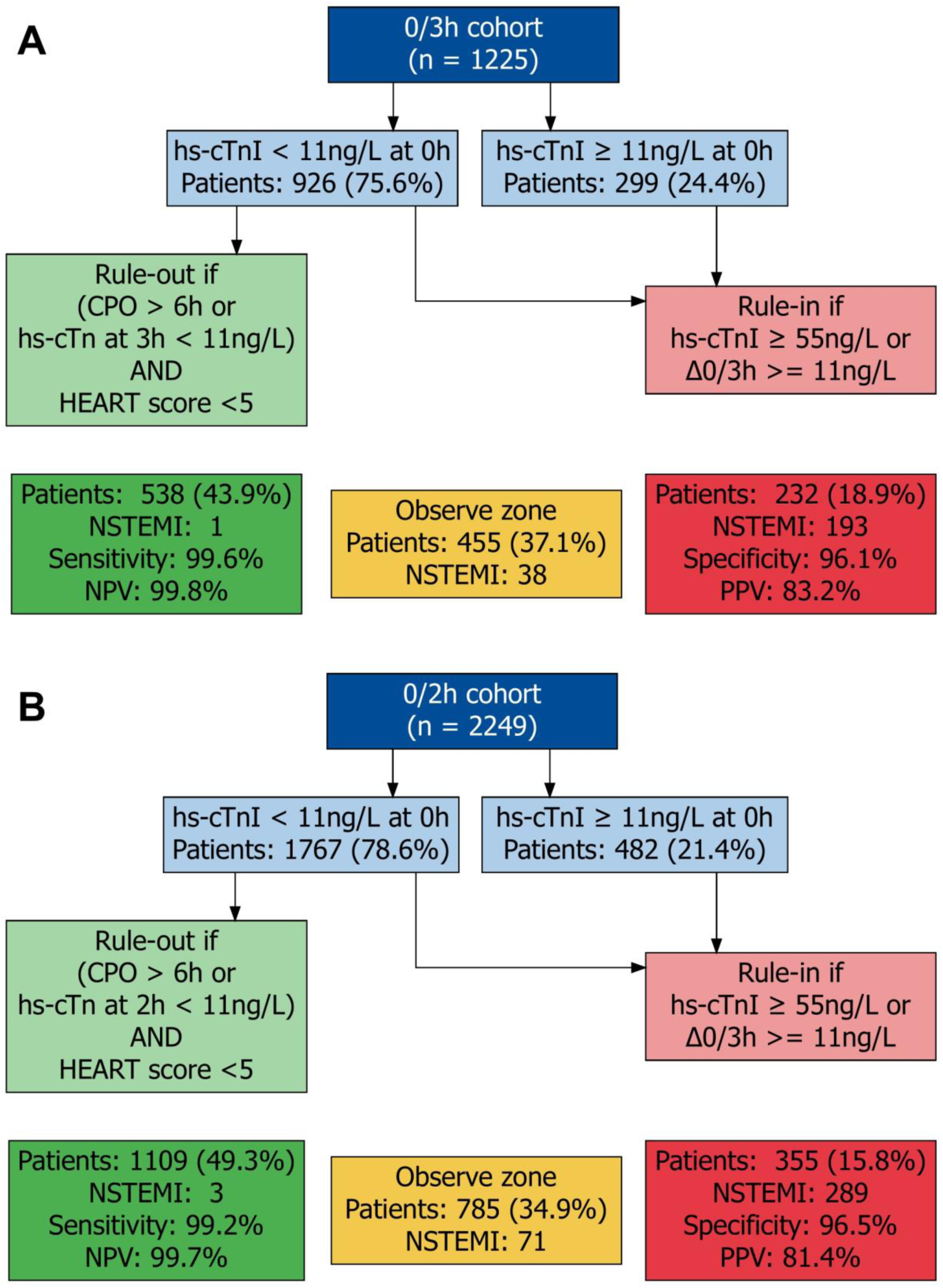
Validation of the complete ESC 0/3h-algorithm using hs-cTnI-VITROS and HEART score. Validation of the ESC 0/3-hour algorithm using hs-cTnI-VITROS with HEART score–based clinical risk stratification irrespective of resolution of symptoms in the (A) 0/3-hour cohort and (B) 0/2-hour cohort. The algorithm displays patient flow and diagnostic safety and efficacy for rule-out, observe, and rule-in categories. Abbreviations: HEART-History, ECG, Age, Risk factors and Troponin Score, hs-cTnI–high-sensitivity cardiac troponin I, NPV–negative predictive value, NSTEMI–non-ST-elevation myocardial infarction, PPV–positive predictive value, Sens–sensitivity, Spec–specificity, URL–upper reference limit.

### Prognosis of complete ESC 0/3h-algorithms using hs-cTnI-VITROS and HEART score

In the main cohort, death from any cause within 5 years occurred in 162 patients, with cumulative mortality rates of 4.9% (25 deaths) in the rule-out group, 16.7% (70 deaths) in the observe zone, and 29.8% (67 deaths) in the rule-in group (**Figure 5A**). The 5-year cumulative incidence for MI or CV death was 3.3% (95%CI: 2–5.2%) in the rule-out group, 14% (95%CI: 11–18%) in the observe zone, and 27% (95%CI: 21–33%) in the rule-in group (**Supplementary Figure 12A**). Sensitivity analysis using the 2h study blood sample revealed similar findings (**Figure 5B and Supplementary Figure 12B**).

**Figure 5.**
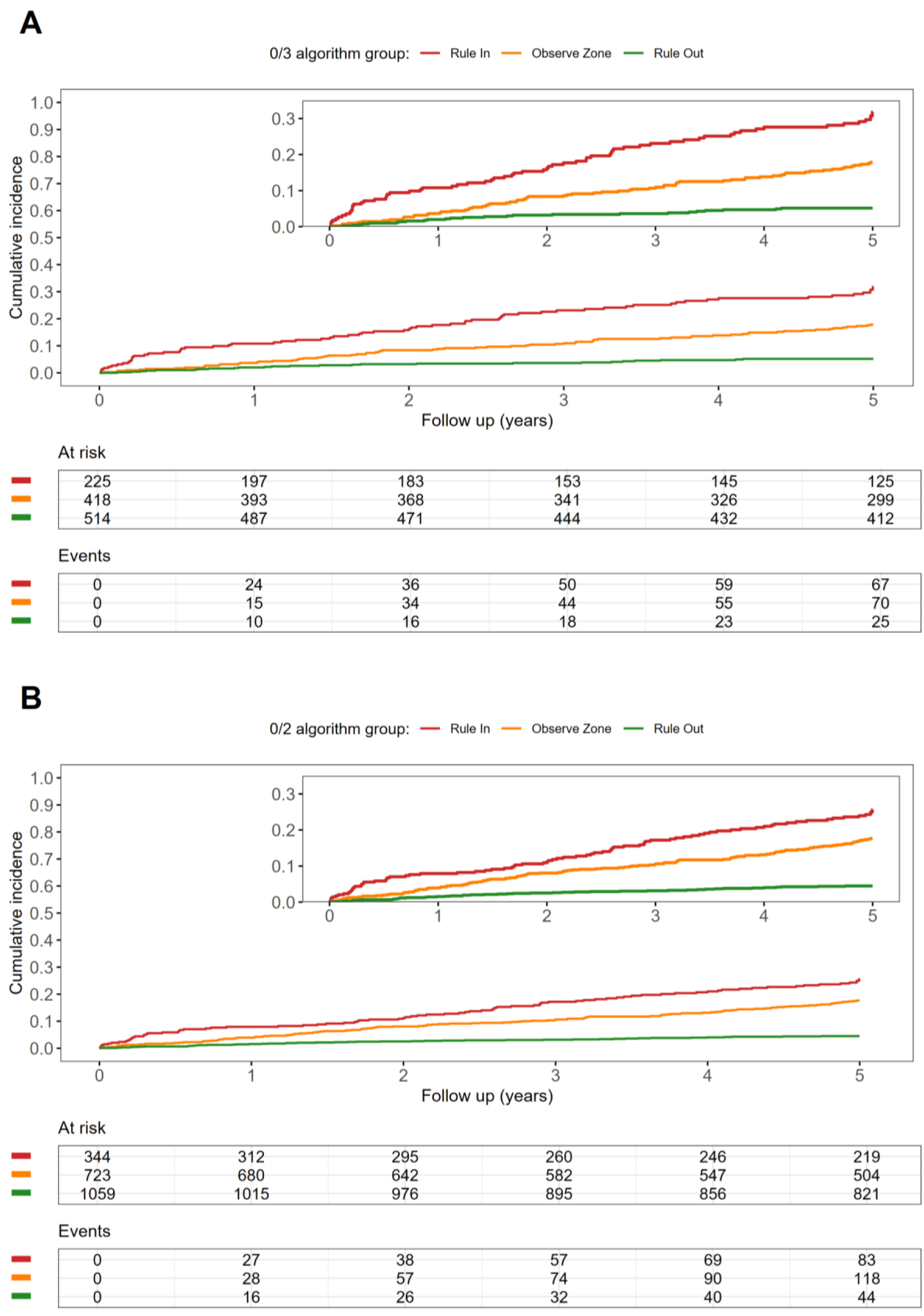
Prognosis of the complete ESC 0/3h-algorithm using hs-cTnI-VITROS and HEART score. Kaplan–Meier plots depicting 5-year cumulative incidence of all-cause death in patients stratified by the complete ESC 0/3-hour algorithm using hs-cTnI-VITROS and HEART score in the (A) 0/3-hour cohort and (B) 0/2-hour cohort. Log-rank test for between-group comparison for all-cause death, p<0.001.

### Validation of simplified ESC 0/3h-algorithms using only hs-cTnI-VITROS

In the main cohort (n=1225; 18.9% with NSTEMI), a URL-based cutoff of <11 ng/L (applied at 0h and 3h, or only at 0h in patients presenting >6h after chest pain onset [direct rule-out]), classified 889 patients (72.6%) as rule-out, yielding a sensitivity of 94.4% and a NPV of 98.5%. URL-based rule-in criteria (hs-cTnI ≥55 ng/L or an absolute 0/3h change ≥11 ng/L) identified 228 patients (18.6%) as rule-in, with a specificity of 96.2% and a PPV of 83.3%. A total of 108 patients (8.8%) remained in the observe zone, of which 29 patients (26.9%) had an adjudicated NSTEMI (**Supplementary Figure 13A**). Sensitivity analysis using the 2h study blood sample revealed similar findings (**Supplementary Figure 13B**).

### Validation of ESC 0/3h-algorithms using LOQ-based absolute change criteria of hs-cTnI-VITROS

In the main cohort (n=1225; 18.9% with NSTEMI), a URL-based cutoff of <11 ng/L (applied at 0h) in combination with a 3h absolute change of ≤1.5 ng/L classified 803 patients (65.6%) as rule-out, resulting in a sensitivity of 97.4% and a NPV of 99.3%. The URL-based rule-in criteria for NSTEMI (hs-cTnI ≥55 ng/L or an absolute 0/3h change ≥11 ng/L) ruled-in 235 patients (19.2%) with a specificity of 95.9% and a PPV of 82.6%. A total of 187 patients (15.3%) remained in the observe zone, of whom 32 patients (17.1%) had an adjudicated NSTEMI (**Supplementary Figure 14A**). Comparable results were found when using LOQ-based percentage change criteria of <20% (**Supplementary Figure 15A**). A modification requiring a 3h absolute change of 0 ng/L classified only 73 patients (6%) as rule-out, resulting in a sensitivity of 100% and a NPV of 100% (**Supplementary Figure 16A**). Sensitivity analysis using the 2h study blood sample revealed similar findings (**Supplementary Figure 14B, Supplementary Figure 15B**, **Supplementary Figure 16B**).

## Discussion

This large, prospective, international multicenter study addressed uncertainties regarding the safety and efficacy of different modifications of the ESC 0/3h-algorithm using the hs-cTnI-VITROS assay. We report seven major findings:

**First**, the complete ESC 0/3h-algorithm combining hs-cTnI-VITROS with clinical risk stratification including the GRACE-score showed very high sensitivity and NPV for the rule-out of NSTEMI. However, very high rule-out performance was achieved at the cost of limited efficacy (30.9%), with approximately half of patients remaining in the observe zone. **Second**, substituting the GRACE score with the HEART score significantly improved the efficacy of the ESC 0/3h-algorithm without compromising safety. The combination of hs-cTnI-VITROS with a HEART score <5 enabled the rule-out of nearly half of patients, while maintaining very high sensitivity and NPV ≥99%. These results were replicated when hs-cTnI-Architect and hs-cTnT-Elecsys Gen-5 were substituted for hs-cTnI-VITROS using their established URL cutoffs, confirming the robustness of HEART score–based risk stratification within the ESC 0/3h-algorithm. **Third,** diagnostic performance of the complete ESC 0/3h-algorithms was consistent across all predefined subgroups and remained robust in sensitivity analyses restricted to the alternatively used endpoint of type 1 NSTEMI.^10,31–33^ **Fourth**, long-term follow-up through 5 years confirmed very low rates of mortality and cardiovascular events among patients triaged to rule-out, supporting the prognostic safety of these strategies. **Fifth**, the simplified ESC 0/3h-algorithm relying exclusively on hs-cTnI-VITROS concentrations did not achieve the safety requirements recommended by current clinical practice guidelines^3,4^. Although a hs-cTnI-only approach had very high efficacy (72.6%), its sensitivity was only 94.4%, mainly due to the high proportion of patients with NSTEMI presenting with an 0h hs-cTn concentration below the URL. **Sixth,** adding LOQ-based absolute change criteria consistent with reporting limits in clinical practice (≤1.5 ng/L) to the simplified ESC 0/3h-algorithms failed to achieve contemporary safety targets for rule-out of NSTEMI. In a subsequent, more restrictive approach, requiring an absolute change of 0 ng/L achieved very high sensitivity and NPV, but at the cost of severely reduced efficacy, triaging only a small proportion of patients as rule-out and leaving the vast majority in the observe zone. Both findings highlight that, in contrast to accelerated the 0/1h- and 0/2h-algorithms^12–14^, URL-based strategies cannot be safely simplified to a purely hs-cTn-driven approach. **Seventh**, sensitivity analyses using the 2h-(rather than the 3h)-study blood sample, which allowed to increase the sample size to 2249 patients, overall revealed consistent findings.

These findings have important clinical implications, particularly given the recent FDA clearance of the hs-cTnI-VITROS assay and the operational simplicity of the ESC 0/3h-algorithm, which make it an attractive option for physicians and institutions. For the first time, physicians and institutions using hs-cTnI-VITROS assay have a detailed and generalizable validation study regarding safety and efficacy for its intended use within different modifications of the ESC 0/3h-algorithm in patients presenting with suspected AMI to the ED. Overall, using the complete algorithm, but replacing the GRACE-with the HEART score seemed to best balance safety and efficacy. Consistent with current ESC and ACC/AHA guideline recommendations, all algorithms must be applied in conjunction with careful clinical assessment and 12-lead ECG to ensure optimal diagnostic performance.^3,4^

These findings extend and corroborate prior studies demonstrating excellent diagnostic performance of hs-cTnI-VITROS when used with accelerated 0/1-hour and 0/2-hour algorithms, with performance comparable to the best validated hs-cTn- assays, including hs-cTnT-Elecsys and hs-cTnI-Architect.^12–14^ Importantly, accelerated 0/1h and 0/2h algorithms are currently preferred where available, while the ESC 0/3h-algorithm is recommended as an alternative diagnostic strategy.^3^ In these settings, our findings provide contemporary evidence that the ESC 0/3h-algorithm can be applied safely and with clinically meaningful efficacy using the hs-cTnI-VITROS assay with established clinical risk scores, consistent with observations by Supples et al. that integration of the HEART score within a URL-based ESC algorithm is required to achieve contemporary safety targets.^34^ Similarly, a study by Chapman et al. reported that combining the ESC 0/3h-algorithm with a low-risk HEART score (<4) increased NPV to 99.7% but reduced the rule-out proportion to ∼25%, whereas in our study a slightly higher threshold (<5) maintained NPV 99.8% while increasing rule-out efficacy to 43.9%. A less restrictive HEART score threshold (<5) can be justified in the ESC 0/3h-algorithm because the previous step requiring URL-based hs-cTn testing already defines a low-risk candidate set. This concept is further supported by large implementation data showing that algorithms incorporating both hs-cTn testing and HEART score–based risk stratification increased early discharges without increasing 30-day death or myocardial infarction.^35^

This study has several strengths. First, it is based on a large prospective, international, multicenter cohort with central adjudication of the final diagnosis by two independent cardiologists, increasing validity and generalizability of results. Second, our findings remained consistent across predefined subgroups and several sensitivity analyses. Third, by incorporating long-term follow-up, this study was able to provide further reassurance regarding the safety of different algorithms, and the appropriateness of the rule-in cutoffs.

Some limitations merit consideration. First, APACE was conducted in patients presenting with acute chest discomfort to the ED; applicability to lower-risk populations, such as those presenting to outpatient clinics, requires further investigation. Second, we cannot generalize these findings to patients with endstage renal disease on chronic dialysis, as they were excluded. Third, external validation in an independent geographically diverse cohort remains warranted. Last, the performance of the ESC 0/3h-algorithm was assessed using central adjudication by two independent cardiologists. However, patient care was not guided by these algorithms but based on local hs-cTnT/I-assays. Therefore, additional prospective studies accompanying the implementation into routine clinical care are still needed.

## Conclusions

This large multicenter study demonstrated that the URL-based ESC 0/3h-algorithms using the hs-cTnI-VITROS assay required integration of clinical risk stratification to achieve guideline-recommended safety for the rule-out of NSTEMI. Among the evaluated approaches, combining hs-cTnI-VITROS with the HEART score reached the highest clinical efficacy while maintaining excellent safety, supporting its use for efficient and safe triage in patients presenting with suspected AMI.

## Data Availability

Due to the sensitive nature of the data, it is only available upon request.

## Non-standard Abbreviations and Acronyms

APACE: Advantageous Predictors of Acute Coronary Syndromes Evaluation
CV: Coefficient of variation
DM: Diabetes mellitus
ED: Emergency Department
ESC: European Society of Cardiology
FDA: U.S. Food and Drug Administration
h: hours
hs-cTn: High-sensitivity cardiac troponin
hs-cTnI: High-sensitivity cardiac troponin I
hs-cTnT: High-sensitivity cardiac troponin T
hs-cTnI-VITROS: High-sensitivity cardiac troponin I assay (VITROS, Ortho Clinical Diagnostics)
LOD: Limit of detection
LOQ: Limit of quantification
MACE: Major adverse cardiovascular events
NSTEMI: Non-ST-elevation myocardial infarction
NPV: Negative predictive value
PPV: Positive predictive value
Q1–Q3: 25th–75th percentiles
ROC: Receiver operating characteristics
STARD: Standards for Reporting of Diagnostic Accuracy Studies
URL: Upper reference limit

## Collaborators

Additional APACE investigators^#^ and Collaborators **to be listed in Pubmed:**

Thomas Nestelberger, MD^a,^; Desiree Wussler, MD^a,^; Maria Rubini Gimenez, MD^a,^; Tobias Zimmermann, MD^a^; Jude Formambuh, MD^a^; Christian Puelacher, MD, PhD^a,^; Jeanne du Fay de Lavallaz, MD, PhD^a,^; Julia Reinhardt, PhD^a,^; Kathrin Meissner, SN^a,^; Katharina Rentsch, PhD^a,^; Ksenia Slankamenac, MDPhD Prof.^b^; Beatriz López, MD^c,d^; Gemma Martinez-Nadal^c,d^; Esther Rodriguez Adrada, MD^c,e^; Arnold von Eckardstein, MD^f^; Damian Kawecki, MD^c,g^; Piotr Muzyk, MD^c,g^; Nicolas Geigy, MD^h^; Stephan Steuer, MD^j^. Angelika Hammerer-Lercher, MD^k^; Andreas Buser, MD^a^

^a^Cardiovascular Research Institute Basel (CRIB) and Department of Cardiology, University Hospital Basel, University of Basel.

^b^Emergency Department, University Hospital Zürich, University of Zürich; Switzerland.

^c^GREAT network; Rome, Italy.

^d^Emergency Department, Hospital Clinic, Barcelona, Catalonia, Spain.

^e^Servicio de Urgencias, Hospital Clínico San Carlos, Madrid, Spain.

^f^Department of Laboratory Medicine, University Hospital Zurich, Switzerland.

^g^2nd Department of Cardiology, School of Medicine with the Division of Dentistry in Zabrze, Medical University of Katowice, Poland.

^h^Emergency Department, Kantonsspital Basel-Land, Switzerland.

^i^Department of Internal Medicine, Kantonsspital Basel-Land, Switzerland.

^j^Emergency Department, Claraspital Basel, Switzerland.

^k^Department of Laboratory Medicine, Hospital Feldkirch, Austria.

## Acknowledgements

We are indebted to the patients who participated in the study and to the emergency department staff as well as the laboratory technicians of all participating sites for their most valuable efforts.

